# Dairy consumption and incident cardiovascular disease: a global analysis

**DOI:** 10.1101/2023.11.14.23298545

**Authors:** Pan Zhuang, Xiaohui Liu, Yin Li, Yang Ao, Yuqi Wu, Hao Ye, Xuzhi Wan, Lange Zhang, Denghui Meng, Yimei Tian, Xiaomei Yu, Fan Zhang, Anli Wang, Yu Zhang, Jingjing Jiao

**Author notes:** **Correspondence to:** Jingjing Jiao, PhD, Department of Nutrition, School of Public Health, Department of Endocrinology, The Second Affiliated Hospital, Zhejiang University School of Medicine, 866 Yuhangtang Road, Hangzhou 310058, Zhejiang, China. Or Yu Zhang, PhD, Department of Gastroenterology, The First Affiliated Hospital, Zhejiang University School of Medicine, Department of Food Science and Nutrition, College of Biosystems Engineering and Food Science, Zhejiang University, 866 Yuhangtang Road, Hangzhou 310058, Zhejiang, China. The two authors equally contributed to the work (Pan Zhuang, Xiaohui Liu).

## Abstract

**BACKGROUND:** The role of dairy products in the primary prevention of cardiovascular disease (CVD) remains highly debated. Our study aimed to comprehensively evaluate the association between dairy consumption and CVD risk in Eastern and Western countries.

**METHODS:** Cohort analyses include 487 212 individuals from the China Kadoorie Biobank (CKB) and 418 895 individuals from the UK Biobank (UKB). Dairy consumption was assessed by validated food frequency questionnaires. We calculated hazard ratios using multivariable Cox proportional-hazards models. The primary outcome was incident CVD, coronary heart disease (CHD) and stroke. An updated meta-analysis of prospective cohort studies was further conducted.

**RESULTS:** A total of 98 954 CVD cases occurred during a mean follow-up of 8.6 years in CKB and 11.3 years in UKB. In CKB, regular dairy consumption (mainly liquid whole milk) was not associated with CVD risk but significantly associated with a 9% (95% confidence interval [CI], 5% to 13%) higher CHD risk and a 6% (95% CI, 3% to 9%) lower stroke risk compared with non/rare consumers. In UKB, total dairy consumption was associated with lower risk of CVD, CHD and ischemic stroke. Cheese consumption was associated with lower CVD risk. Multivariable-adjusted hazard ratios (HRs) (95% CIs) comparing ≥ 7 times/week to the < 2 times/week of cheese were 0.88 (0.83–0.94) for CVD, 0.88 (0.82–0.94) for CHD, and 0.97 (0.85–1.11) for stroke. Semi-skimmed milk consumers had decreased risk of CVD and stroke. In the updated meta-analysis, total dairy consumption was significantly associated with a lower risk of CVD (relative risk [RR], 0.963; 95% CI, 0.932 to 0.995; 26 risk estimates) and stroke (RR, 0.94; 95% CI, 0.90 to 0.98; 14 risk estimates). Inverse associations with CVD incidence were found for cheese (0.94; 0.91 to 0.97; 20 risk estimates) and low-fat dairy (0.96; 0.92 to 0.99; 20 risk estimates) but not milk and yogurt.

**CONCLUSIONS:** Total dairy consumption is associated with a lower risk of total CVD and stroke overall but relationships vary by types of dairy products. Cheese and low-fat dairy consumption may be advocated for the primary prevention of CVD.

**Clinical Perspective:** *What Is New?:*

- Whether dairy products are protective for cardiovascular disease (CVD) remains highly debated.
- This global largest analysis that included two original cohorts of 0.9 million participants from China and the UK and an updated meta-analysis demonstrates that higher consumption of total dairy products was associated with lower risk of CVD and stroke overall.
- For dairy subtypes, cheese and low-fat dairy consumption was associated with lower CVD risk while no significant association was observed for milk, yogurt, and high-fat dairy consumption.

*What Are the Clinical Implications?:*

- Results from our two large cohort studies and updated meta-analysis support that dairy product consumption is protective for CVD, especially stroke, and provide compelling evidence relevant to dietary guidelines.
- Considering specific dairy subtypes, cheese, and low-fat dairy products may be protective and should be advocated for CVD prevention.

---

Cardiovascular disease (CVD) is the largest contributor to death globally.^1^ Adopting healthy dietary patterns is one of the cornerstones of primary prevention of CVD. Thereinto, although dairy consumption features in many dietary guidelines, its role in a heart-healthy diet remains highly debated.^2^ Dairy products contain various beneficial nutrients, including high biological value protein, milk fat globule phospholipids, and vitamins and minerals that could improve CVD risk factors,^3–5^ whereas saturated fats^6^ and multiple anabolic hormones^7^ in dairy products might adversely affect the health benefit. Previous prospective studies linking dairy consumption with CVD outcomes have yielded conflicting results. Some cohort studies reported a protective relationship between dairy consumption and CVD outcomes,^8–12^ whereas others showed no significant associations^13–16^ or even positive associations.^17, 18^ Meta-analyses also yielded inconsistent conclusions on associations of dairy intake with CHD and stroke risk.^19, 20^ Notably, heterogeneity between included studies was considerable and the overall quality of the evidence was low to moderate.

Prevailing recommendations advocate low-fat or non-fat dairy over whole-fat dairy.^21^ However, scientific evidence for this recommendation was scant and inconsistent.^19^ Importantly, different subtypes of dairy products may confer divergent health effects after processing. Fermented milk products such as yogurt contain probiotics that can favorably regulate gut microbiome,^22^ whereas cheese is rich in sodium which may elevate blood pressure when consumed in excessive amounts.^23^ Nonetheless, cheese is also a fermented food that can contain vitamin K2, high levels of milk fat globule membrane, as well as probiotics. Furthermore, previous epidemiological studies were largely conducted in western countries, where the consumption level of dairy products especially cheese is high and usually correlated with a higher socio-economic position.^24, 25^ In Asia where strokes are more common than CHD, only few studies demonstrated an inverse association of dairy consumption with stroke.^8, 26^ Overall, evidence from large cohort studies in both western and non-western countries is needed to make global policy recommendations.

To address the above-mentioned gaps in knowledge, we followed 0.9 million individuals from the UK Biobank (UKB) study and the China Kadoorie Biobank (CKB) study to evaluate the associations of dairy product consumption with incident CVD, CHD and stroke. We also performed an updated systematic review of the literature and meta-analysis of dairy product intake and incident CVD risk which included our findings to address the role of dairy consumption in CVD prevention and improve dietary guidelines.

## METHODS

### Study Population

CKB is one of the largest cohort studies that recruited over 500,000 adults from ten geographically diverse areas across China during 2004-2008.^27^ The CKB study received ethical approval from the Oxford University Tropical Research Ethics Committee, the Chinese Centre for Disease Control and Prevention (CDC) Ethical Review Committee and the local CDC of each study area. All participants gave written informed consent. For this analysis, participants with a history of CVD or cancer were excluded at baseline, which resulted in a sample of 487 212 individuals in the CKB.

UKB is also a large prospective study of more than 500,000 people who were aged 37–73 years and recruited from one of 22 assessment centers across the UK between 2007 and 2010.^28^ The UK Biobank received ethical approval from the research ethics committee (REC reference for UK Biobank 11/NW/0382). Among 502 476 participants, we excluded participants with a history of CVD or cancer at baseline and participants who withdrew during the follow-up (data cannot be used).

Furthermore, we excluded persons without data on cheese consumption frequency from the food frequency questionnaire (FFQ) or those without information about 24-hour dietary recalls. Finally, 418 895 individuals in the UKB remained in the final analytical samples for cheese consumption and 183 446 individuals remained for individual dairy products. The detailed flow chart is shown in Figure S1.

### Dietary Assessments

In the CKB, participants were asked about the consumption frequency of 12 major food groups, including total dairy products over the preceding year by a qualitative FFQ. The weighted kappa statistics of dairy consumption frequency were 0.82 for reproducibility and 0.75 for validity, comparing two FFQs conducted in the second and third surveys with the baseline FFQ, which implicated good performance of the FFQ^29^. Subtypes of dairy products were not included in the baseline FFQ and thus were not analyzed in CKB.

In the UKB, participants completed a touch to Screen short FFQ that consisted of 29 diet questions over the past 12 months, including frequency of cheese intake (0, <1, 1, 2 to 4, 5 to 6, ≥7 times a week) and type of milk (never/rarely have milk, full, semi-skimmed, skimmed cream, soya, other) in which they could select multiple types of milk they drank. Besides, participants were invited to complete a 24-hour dietary questionnaire that inquired about the consumption of nearly 200 foods and drinks including various dairy products (milk, cheese, yogurt and ice cream). Five separate occasions of 24-hour dietary recalls were conducted during 2011-2012 to provide an average measure for individuals (repeated measurement per person). The dietary touch-screen FFQ has been validated against online 24-hour dietary assessments^30^ and the latter has been validated using biomarkers reported elsewhere.^31^

### Ascertainment of Incident Cardiovascular Disease

Detailed information used to define incident CVD cases including fatal or non-fatal CHD and stroke is presented in Table S1. Incident cases of CVD were identified by using linkages with disease registries and national health insurance claim databases and further supplemented with annual active follow-up in CKB. In UKB, information on the CVD cases of all participants was obtained from cumulative hospital inpatient records. All events were ascertained using the International Classification of Diseases, 10th Revision (ICD–10).

### Statistical Analysis

The main exposures of interest were the frequency of total dairy consumption in CKB and the frequency of cheese intake (<2, 2 to 4, 5 to 6, or ≥7 times a week), milk type, and total dairy consumption in UKB. The intake of dairy products (0, ≤0.5, 0.5 to 1, or >1 serving per day), including milk, yogurt, and cheese (0, ≤0.5, or >0.5 serving per day), was categorized into predefined categories based on consumption distributions.

The person-year was calculated from the date of entry to the time of CVD diagnosis, lost to follow-up, death, or the end date of follow-up (December 31, 2016 for CKB, and 31 December 2020 for UKB), whichever occurred earlier. Only 1.2% of individuals in CKB and 0.3% in UKB were lost to follow-up and censored in analyses. Cox proportional hazards regression model was used to estimate hazard ratios (HRs) and 95% confidence intervals (CIs) of CVD risk for total or each type of dairy product consumption after checking the violation of the proportional hazard assumption. To control known and potential confounders, multivariable models were sequentially adjusted for age, sex, race, study area (for CKB)/assessment centers (for UKB), body mass index (BMI), education level, household income, Townsend deprivation index (TDI, only in UKB), smoking status, alcohol drinking, physical activity, history of hypertension, history of diabetes, family history of CVD, use of vitamins, minerals and aspirin, and consumption frequency of red meat, processed red meat (only in UKB), fish, oily fish (only in UKB), non-oily fish (only in UKB), poultry, vegetables, fruits, and eggs (all categories of consumption). All missing data were coded as an independent category if necessary. The linear trend was tested by fitting the ordinal dairy variables as continuous variables in the models.

As dairy products are one of the major sources of dietary protein, we used substitution analysis to estimate the theoretical effect on CVD risk of substituting one serving of dairy products for an equivalent serving of other common alternative protein sources, including red/processed meat, fish, poultry, eggs, and soybean/legumes.^11^ We further examined whether the documented associations varied by subgroups according to baseline characteristics, including age, sex, BMI, household income, smoking status, alcohol intake frequency, physical activity, diet quality, hypertension, diabetes, and family history of CVD. Besides, we conducted several sensitivity analyses. First, we adjusted a healthy diet score^32^ ^33^ to evaluate the influence of the overall diet quality. Second, lipid-lowering drugs or anti-hypertensive medications were further adjusted in the model. Third, we further excluded incident CVD cases within the first 2 years of follow-up or participants with extreme BMIs (<18.5 or >40 kg/m^2^). Finally, participants were censored at a 5-y follow-up. In addition, in CKB analysis, we used a multivariable Cox frailty model with random intercepts to account for center clustering (10 regions). In UKB analysis, we further adjusted for energy intake or salt added to food to see whether the main findings altered.

All statistical analyses were conducted with SAS 9.4 (SAS Institute, Cary, NC, USA) and a two-sided P < 0.05 was considered statistically significant.

### Meta-analysis

We performed a systematic review and updated meta-analysis including UKB and CKB studies as well as previous prospective cohort studies which explored the relationship of dairy product intake with CVD risk in the general population. Table S2 shows the search strategy. Additional details of the meta-analysis are provided (for details, see the Expanded Methods in the Supplemental Material).

## RESULTS

### Cohort Analyses

During a follow-up of 4 190 676 person-years in CKB and 4 736 113 person-years in UKB, we documented 66 132 CVD cases in CKB and 32 822 CVD cases in UKB. In CKB, participants who consumed dairy products more frequently tended to be women, higher-educated, high-income class, urban residents and vitamin and mineral supplements users, have diabetes and family history of CVD, and consume fruits and eggs more frequently (Table S3). In UKB, individuals with higher total dairy consumption were more likely to exercise, be more educated, take vitamin and mineral supplements, and consume oily fish and fruits more frequently, whereas they drank alcohol less frequently and had a lower hypertension prevalence (Table S4). Characteristics of participants by cheese consumption (the main subtype of dairy in UKB) and milk types in UKB are shown in Tables S5 and S6.

Compared to non/rare consumers, those who consumed at least 4 times/week of dairy had no significant association with CVD after the multivariable adjustment in CKB (HR 1.09, 95% CI 0.97–1.03, P-trend=0.470). Regular dairy consumption was related to a 9% higher risk of CHD (HR 1.09, 95% CI 1.05–1.13, P-trend<0.001) but a 6% lower risk of stroke (HR 0.94, 95% CI 0.91–0.97, P-trend=0.005), especially hemorrhagic stroke (HR 0.76, 95% CI 0.69–0.83, P-trend<0.001) (**Table 1**). In UKB, total dairy intake was inversely associated with incident CVD (HR 0.93, 95% CI 0.88– 0.98, P-trend=0.004), CHD (HR 0.93, 95% CI 0.88–0.99, P-trend=0.014), and ischemic stroke (HR 0.86, 95% CI 0.75–0.99, P-trend=0.036) (**Table 2**). For individual dairy products, cheese consumption was associated with lower CVD and CHD risk.

**Table 1.**
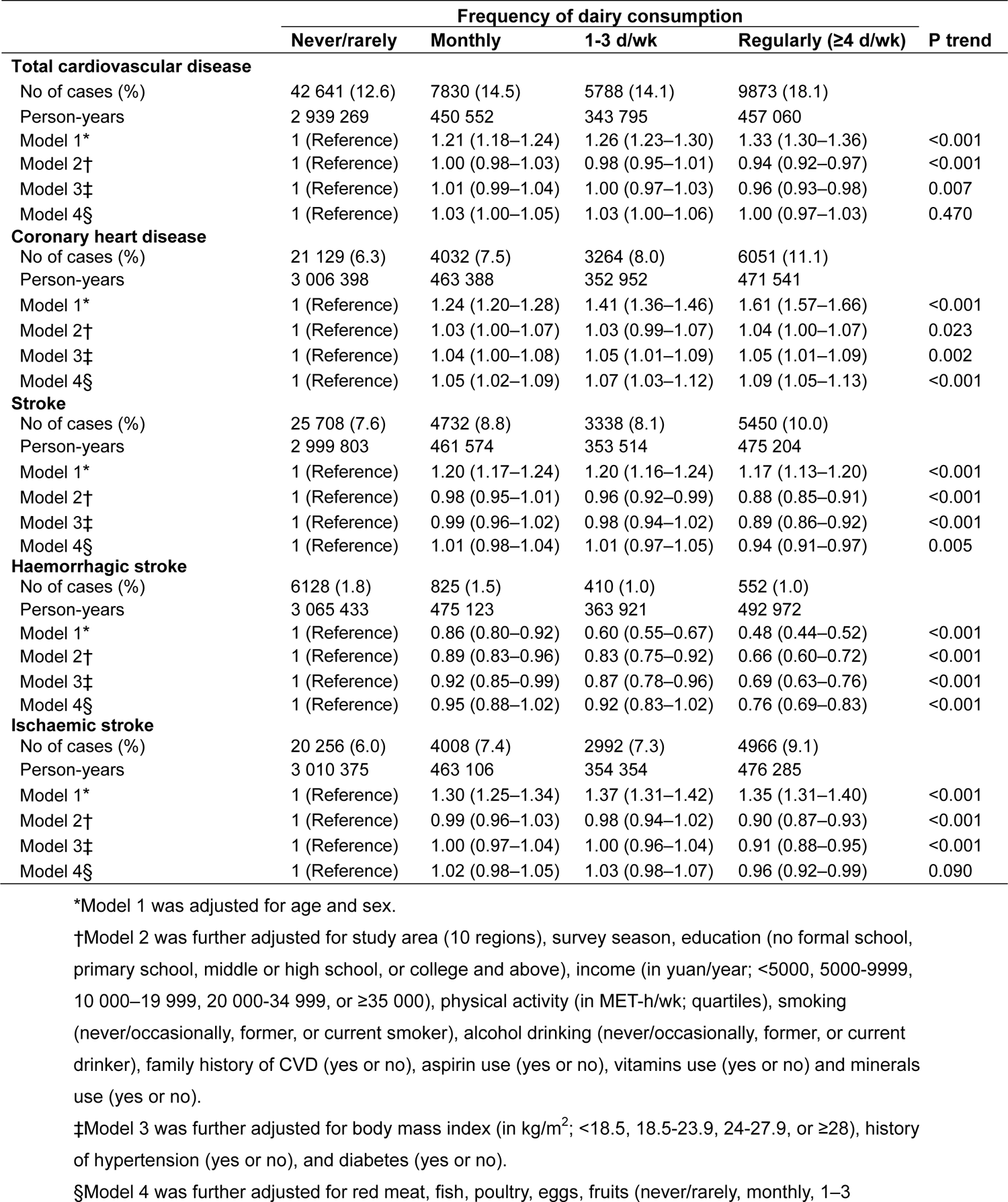
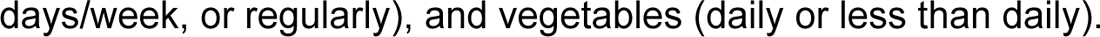
Hazard ratios (95% confidence intervals) for incident cardiovascular disease according to categories of dairy consumption in China Kadoorie Biobank days/week, or regularly), and vegetables (daily or less than daily).

|  | Frequency of dairy consumption |  |  |  | P trend |
| --- | --- | --- | --- | --- | --- |
|  | Never/rarely | Monthly | 1-3 d/wk | Regularly (≥4 d/wk) |  |
| **Total cardiovascular disease** |  |  |  |  |  |
| No of cases (%) | 42 641 (12.6) | 7830 (14.5) | 5788 (14.1) | 9873 (18.1) |  |
| Person-years | 2 939 269 | 450 552 | 343 795 | 457 060 |  |
| Model 1* | 1 (Reference) | 1.21 (1.18–1.24) | 1.26 (1.23–1.30) | 1.33 (1.30–1.36) | <0.001 |
| Model 2† | 1 (Reference) | 1.00 (0.98–1.03) | 0.98 (0.95–1.01) | 0.94 (0.92–0.97) | <0.001 |
| Model 3‡ | 1 (Reference) | 1.01 (0.99–1.04) | 1.00 (0.97–1.03) | 0.96 (0.93–0.98) | 0.007 |
| Model 4§ | 1 (Reference) | 1.03 (1.00–1.05) | 1.03 (1.00–1.06) | 1.00 (0.97–1.03) | 0.470 |
| **Coronary heart disease** |  |  |  |  |  |
| No of cases (%) | 21 129 (6.3) | 4032 (7.5) | 3264 (8.0) | 6051 (11.1) |  |
| Person-years | 3 006 398 | 463 388 | 352 952 | 471 541 |  |
| Model 1* | 1 (Reference) | 1.24 (1.20–1.28) | 1.41 (1.36–1.46) | 1.61 (1.57–1.66) | <0.001 |
| Model 2† | 1 (Reference) | 1.03 (1.00–1.07) | 1.03 (0.99–1.07) | 1.04 (1.00–1.07) | 0.023 |
| Model 3‡ | 1 (Reference) | 1.04 (1.00–1.08) | 1.05 (1.01–1.09) | 1.05 (1.01–1.09) | 0.002 |
| Model 4§ | 1 (Reference) | 1.05 (1.02–1.09) | 1.07 (1.03–1.12) | 1.09 (1.05–1.13) | <0.001 |
| **Stroke** |  |  |  |  |  |
| No of cases (%) | 25 708 (7.6) | 4732 (8.8) | 3338 (8.1) | 5450 (10.0) |  |
| Person-years | 2 999 803 | 461 574 | 353 514 | 475 204 |  |
| Model 1* | 1 (Reference) | 1.20 (1.17–1.24) | 1.20 (1.16–1.24) | 1.17 (1.13–1.20) | <0.001 |
| Model 2† | 1 (Reference) | 0.98 (0.95–1.01) | 0.96 (0.92–0.99) | 0.88 (0.85–0.91) | <0.001 |
| Model 3‡ | 1 (Reference) | 0.99 (0.96–1.02) | 0.98 (0.94–1.02) | 0.89 (0.86–0.92) | <0.001 |
| Model 4§ | 1 (Reference) | 1.01 (0.98–1.04) | 1.01 (0.97–1.05) | 0.94 (0.91–0.97) | 0.005 |
| **Haemorrhagic stroke** |  |  |  |  |  |
| No of cases (%) | 6128 (1.8) | 825 (1.5) | 410 (1.0) | 552 (1.0) |  |
| Person-years | 3 065 433 | 475 123 | 363 921 | 492 972 |  |
| Model 1* | 1 (Reference) | 0.86 (0.80–0.92) | 0.60 (0.55–0.67) | 0.48 (0.44–0.52) | <0.001 |
| Model 2† | 1 (Reference) | 0.89 (0.83–0.96) | 0.83 (0.75–0.92) | 0.66 (0.60–0.72) | <0.001 |
| Model 3‡ | 1 (Reference) | 0.92 (0.85–0.99) | 0.87 (0.78–0.96) | 0.69 (0.63–0.76) | <0.001 |
| Model 4§ | 1 (Reference) | 0.95 (0.88–1.02) | 0.92 (0.83–1.02) | 0.76 (0.69–0.83) | <0.001 |
| **Ischaemic stroke** |  |  |  |  |  |
| No of cases (%) | 20 256 (6.0) | 4008 (7.4) | 2992 (7.3) | 4966 (9.1) |  |
| Person-years | 3 010 375 | 463 106 | 354 354 | 476 285 |  |
| Model 1* | 1 (Reference) | 1.30 (1.25–1.34) | 1.37 (1.31–1.42) | 1.35 (1.31–1.40) | <0.001 |
| Model 2† | 1 (Reference) | 0.99 (0.96–1.03) | 0.98 (0.94–1.02) | 0.90 (0.87–0.93) | <0.001 |
| Model 3‡ | 1 (Reference) | 1.00 (0.97–1.04) | 1.00 (0.96–1.04) | 0.91 (0.88–0.95) | <0.001 |
| Model 4§ | 1 (Reference) | 1.02 (0.98–1.05) | 1.03 (0.98–1.07) | 0.96 (0.92–0.99) | 0.090 |

\*Model 1 was adjusted for age and sex.
†Model 2 was further adjusted for study area (10 regions), survey season, education (no formal school, primary school, middle or high school, or college and above), income (in yuan/year; <5000, 5000-9999, 10 000–19 999, 20 000-34 999, or $\geq 35$ 000), physical activity (in MET-h/wk; quartiles), smoking (never/occasionally, former, or current smoker), alcohol drinking (never/occasionally, former, or current drinker), family history of CVD (yes or no), aspirin use (yes or no), vitamins use (yes or no) and minerals use (yes or no).
‡Model 3 was further adjusted for body mass index (in kg/m<sup>2</sup>; <18.5, 18.5-23.9, 24-27.9, or $\geq 28$ ), history of hypertension (yes or no), and diabetes (yes or no).
§Model 4 was further adjusted for red meat, fish, poultry, eggs, fruits (never/rarely, monthly, 1–3

**Table 2.**
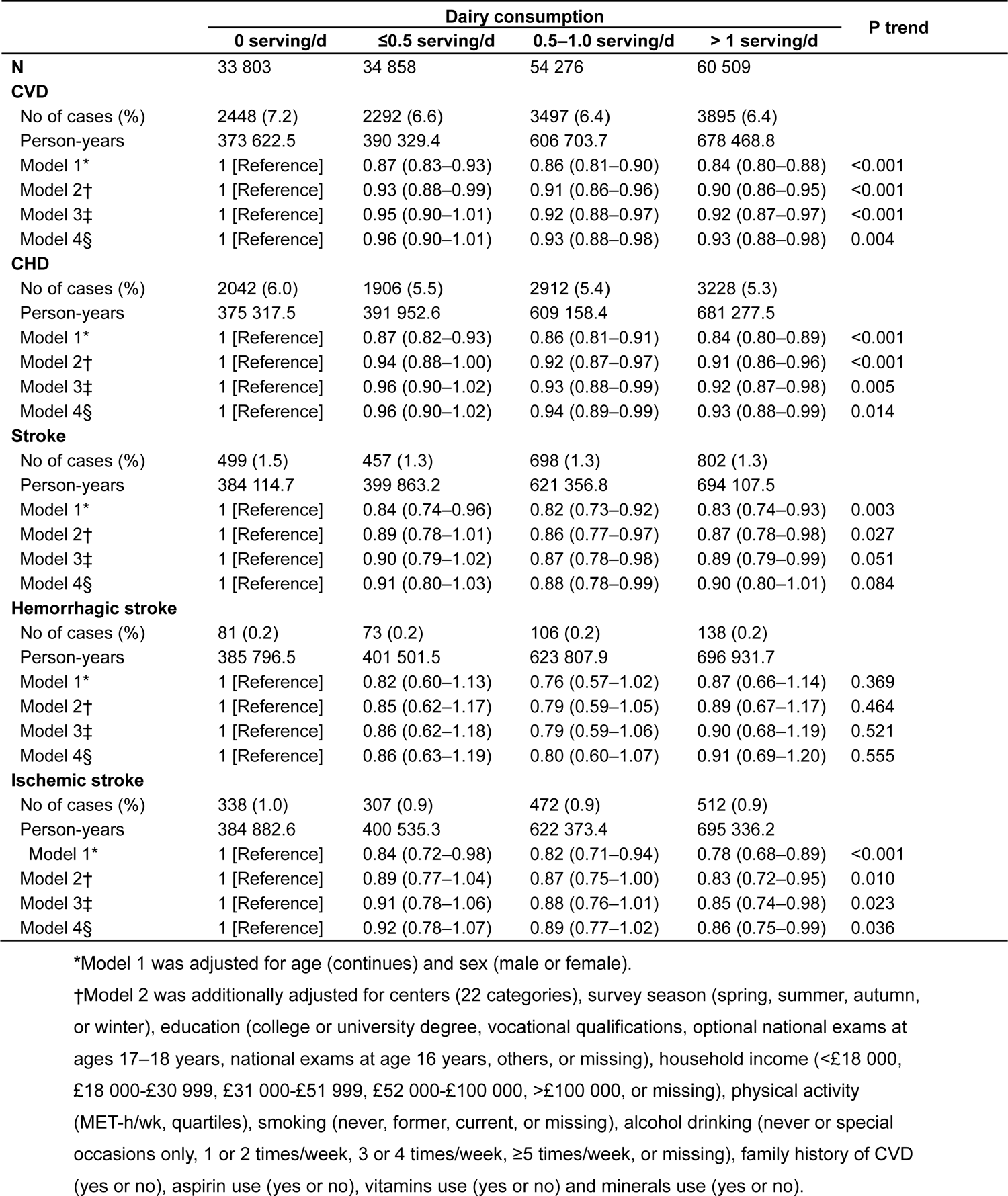
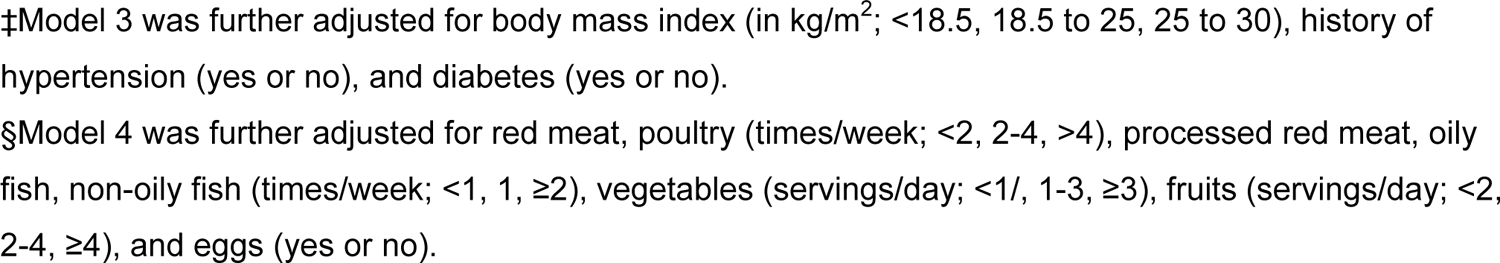
Hazard ratios (95% confidence intervals) for incident cardiovascular disease according to categories of dairy consumption in UK Biobank.

|  | Dairy consumption |  |  |  | P trend |
| --- | --- | --- | --- | --- | --- |
|  | 0 serving/d | ≤0.5 serving/d | 0.5–1.0 serving/d | > 1 serving/d |  |
| **N** | 33 803 | 34 858 | 54 276 | 60 509 |  |
| **CVD** |  |  |  |  |  |
| No of cases (%) | 2448 (7.2) | 2292 (6.6) | 3497 (6.4) | 3895 (6.4) |  |
| Person-years | 373 622.5 | 390 329.4 | 606 703.7 | 678 468.8 |  |
| Model 1* | 1 [Reference] | 0.87 (0.83–0.93) | 0.86 (0.81–0.90) | 0.84 (0.80–0.88) | <0.001 |
| Model 2† | 1 [Reference] | 0.93 (0.88–0.99) | 0.91 (0.86–0.96) | 0.90 (0.86–0.95) | <0.001 |
| Model 3‡ | 1 [Reference] | 0.95 (0.90–1.01) | 0.92 (0.88–0.97) | 0.92 (0.87–0.97) | <0.001 |
| Model 4§ | 1 [Reference] | 0.96 (0.90–1.01) | 0.93 (0.88–0.98) | 0.93 (0.88–0.98) | 0.004 |
| **CHD** |  |  |  |  |  |
| No of cases (%) | 2042 (6.0) | 1906 (5.5) | 2912 (5.4) | 3228 (5.3) |  |
| Person-years | 375 317.5 | 391 952.6 | 609 158.4 | 681 277.5 |  |
| Model 1* | 1 [Reference] | 0.87 (0.82–0.93) | 0.86 (0.81–0.91) | 0.84 (0.80–0.89) | <0.001 |
| Model 2† | 1 [Reference] | 0.94 (0.88–1.00) | 0.92 (0.87–0.97) | 0.91 (0.86–0.96) | <0.001 |
| Model 3‡ | 1 [Reference] | 0.96 (0.90–1.02) | 0.93 (0.88–0.99) | 0.92 (0.87–0.98) | 0.005 |
| Model 4§ | 1 [Reference] | 0.96 (0.90–1.02) | 0.94 (0.89–0.99) | 0.93 (0.88–0.99) | 0.014 |
| **Stroke** |  |  |  |  |  |
| No of cases (%) | 499 (1.5) | 457 (1.3) | 698 (1.3) | 802 (1.3) |  |
| Person-years | 384 114.7 | 399 863.2 | 621 356.8 | 694 107.5 |  |
| Model 1* | 1 [Reference] | 0.84 (0.74–0.96) | 0.82 (0.73–0.92) | 0.83 (0.74–0.93) | 0.003 |
| Model 2† | 1 [Reference] | 0.89 (0.78–1.01) | 0.86 (0.77–0.97) | 0.87 (0.78–0.98) | 0.027 |
| Model 3‡ | 1 [Reference] | 0.90 (0.79–1.02) | 0.87 (0.78–0.98) | 0.89 (0.79–0.99) | 0.051 |
| Model 4§ | 1 [Reference] | 0.91 (0.80–1.03) | 0.88 (0.78–0.99) | 0.90 (0.80–1.01) | 0.084 |
| **Hemorrhagic stroke** |  |  |  |  |  |
| No of cases (%) | 81 (0.2) | 73 (0.2) | 106 (0.2) | 138 (0.2) |  |
| Person-years | 385 796.5 | 401 501.5 | 623 807.9 | 696 931.7 |  |
| Model 1* | 1 [Reference] | 0.82 (0.60–1.13) | 0.76 (0.57–1.02) | 0.87 (0.66–1.14) | 0.369 |
| Model 2† | 1 [Reference] | 0.85 (0.62–1.17) | 0.79 (0.59–1.05) | 0.89 (0.67–1.17) | 0.464 |
| Model 3‡ | 1 [Reference] | 0.86 (0.62–1.18) | 0.79 (0.59–1.06) | 0.90 (0.68–1.19) | 0.521 |
| Model 4§ | 1 [Reference] | 0.86 (0.63–1.19) | 0.80 (0.60–1.07) | 0.91 (0.69–1.20) | 0.555 |
| **Ischemic stroke** |  |  |  |  |  |
| No of cases (%) | 338 (1.0) | 307 (0.9) | 472 (0.9) | 512 (0.9) |  |
| Person-years | 384 882.6 | 400 535.3 | 622 373.4 | 695 336.2 |  |
| Model 1* | 1 [Reference] | 0.84 (0.72–0.98) | 0.82 (0.71–0.94) | 0.78 (0.68–0.89) | <0.001 |
| Model 2† | 1 [Reference] | 0.89 (0.77–1.04) | 0.87 (0.75–1.00) | 0.83 (0.72–0.95) | 0.010 |
| Model 3‡ | 1 [Reference] | 0.91 (0.78–1.06) | 0.88 (0.76–1.01) | 0.85 (0.74–0.98) | 0.023 |
| Model 4§ | 1 [Reference] | 0.92 (0.78–1.07) | 0.89 (0.77–1.02) | 0.86 (0.75–0.99) | 0.036 |

\*Model 1 was adjusted for age (continues) and sex (male or female).
†Model 2 was additionally adjusted for centers (22 categories), survey season (spring, summer, autumn, or winter), education (college or university degree, vocational qualifications, optional national exams at ages 17–18 years, national exams at age 16 years, others, or missing), household income (<£18 000, £18 000–£30 999, £31 000–£51 999, £52 000–£100 000, >£100 000, or missing), physical activity (MET-h/wk, quartiles), smoking (never, former, current, or missing), alcohol drinking (never or special occasions only, 1 or 2 times/week, 3 or 4 times/week, ≥5 times/week, or missing), family history of CVD (yes or no), aspirin use (yes or no), vitamins use (yes or no) and minerals use (yes or no).
‡Model 3 was further adjusted for body mass index (in kg/m<sup>2</sup>; <18.5, 18.5 to 25, 25 to 30), history of hypertension (yes or no), and diabetes (yes or no).
§Model 4 was further adjusted for red meat, poultry (times/week; <2, 2-4, >4), processed red meat, oily fish, non-oily fish (times/week; <1, 1, ≥2), vegetables (servings/day; <1/, 1-3, ≥3), fruits (servings/day; <2, 2-4, ≥4), and eggs (yes or no).

The HRs (95% CIs) comparing the frequency at least 7 times/week of cheese with less than 2 times/week were 0.88 (0.83–0.94) for CVD, 0.88 (0.82–0.94) for CHD, and 0.97 (0.85–1.11) for stroke in the fully adjusted model (Table S7), which was similar to the results from 24 h dietary recalls (Table S8). For subtypes of cheese, both hard cheese and fresh cheese (>0.5 serving/d) were associated with a lower risk of CVD and CHD (Tables S9 to S11). We found milk consumption (>0 to 0.5 serving/d) was associated with a lower risk of hemorrhagic stroke (HR 0.43, 95% CI 0.21–0.87) and yogurt consumption (>0.5 serving/d) was related to decreased ischemic stroke risk (HR 0.86, 95% CI 0.77–0.98), compared with non-consumers (Tables S12 and S13). With regard to different types of milk, compared with participants who never or rarely drank milk, both semi-skimmed and skimmed milk consumers had decreased CVD risk (semi-skimmed: HR 0.92, 95% CI 0.87–0.98; skimmed: HR 0.91, 95% CI 0.86–0.97) and stroke risk (semi-skimmed: HR 0.80, 95% CI 0.71–0.90; skimmed: HR 0.76, 95% CI 0.66–0.86) (Table S14).

We also found significant interactions by sex and history of diabetes in the subgroup analyses in both CKB and UKB (Tables S15 to S18). Moreover, our results did not alter substantially in sensitivity analyses (Tables S19 to S22). In hypothetical substitution analyses (Figure S2), though we didn’t find a significant association for CVD risk when replacing red meats, fish, poultry, or soybeans with dairy products, replacing eggs with dairy products was related to a 12% higher risk of CVD (P<0.001) in CKB. Besides, a higher risk of CHD (P<0.001) and stroke (P=0.039) was also observed when eggs were replaced by total dairy products. Substituting one serving of dairy products for red meats or soybeans was associated with lower stroke risk (HR for red meats 0.93, 95% CI 0.89–0.98; HR for soybeans 0.92, 95% CI 0.87–0.97). No significant results were found for hypothetical substitution analyses in UKB.

### Systematic Review and Meta-Analysis

Overall, 30 publications from 25 prospective cohorts and our results from CKB and UKB were kept in our final meta-analysis (Figure S3, Tables S23 and S24). During a range of 5.5 to 30.0 follow-up years, 73 193 CVD cases were documented among 1 288 420 participants from 30 countries or territories around the world in the previous studies (30 studies) (Table S25).

Although no significant association was found between total dairy intake and incident CVD in the meta-analysis of previously published studies (RR 0.963, 95% CI 0.926–1.001, n=24 risk estimates), the summary RR (95% CI) turned into 0.963 (0.932–0.995) when the results of CKB and UKB studies were added (**Figure 1**).

**Figure 1.**
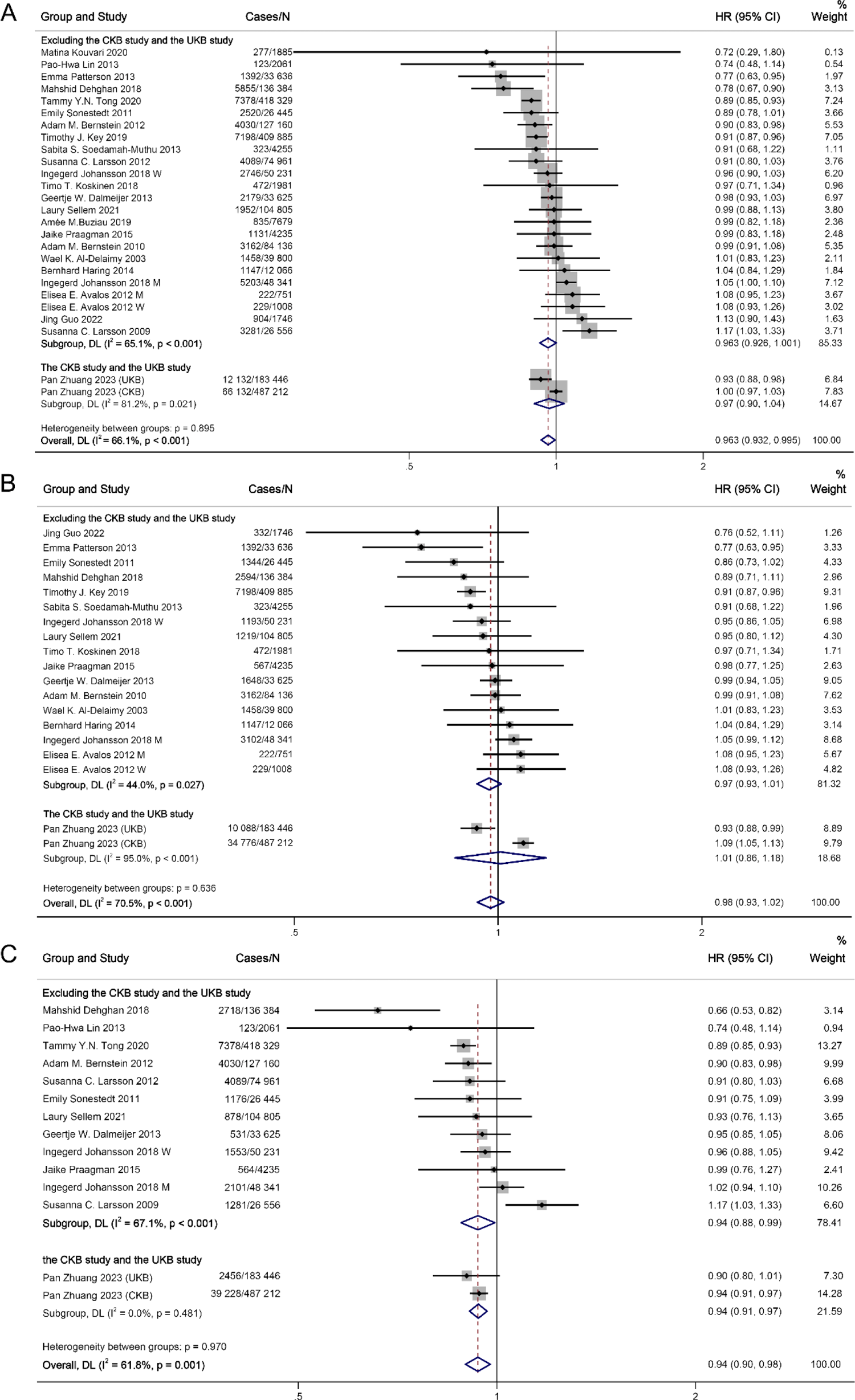
Associations of dairy consumption with cardiovascular disease, coronary heart disease, and stroke risk for high compared with low category of intake using random effects meta-analysis. (A) Cardiovascular disease. (B) Coronary heart disease. (C) Stroke. Squares represent study to Specific relative risk. Gray square areas are proportional to the individual study weight for the overall meta-analysis. Horizontal lines denote 95% CIs. I^2^ refers to the proportion of heterogeneity among studies. M, men; W, women; CKB, China Kadoorie Biobank; UKB, UK Biobank.

Each serving/day increment of total dairy products was related to a 2% lower CVD risk (RR 0.98, 95% CI 0.96–0.99, P<0.001, n=17 risk estimates) (Figure S4). A similar inverse relationship for CVD was also shown in non-linear analysis (P-nonlinear = 0.002, n=12 studies, Figure S5). For subtypes of CVD, the meta-analysis showed dairy consumption had an inverse relationship with total stroke risk (RR 0.94, 95% CI 0.90–0.98, 14 risk estimates, I^2^=61.8%) but null association with CHD risk (RR 0.98, 95% CI 0.93–1.02, 19 risk estimates, I^2^=70.5%, **Figure 1**).

For major subtypes of dairy products, high intake of fermented dairy products, especially cheese, had a protective association with CVD risk (RR for fermented dairy 0.96, 95% CI: 0.94–0.98, n=24 risk estimates; RR for cheese 0.94, 95% CI: 0.91– 0.97, n=20 risk estimates), but not yogurt (RR 0.99, 95% CI 0.93–1.06, n=14 risk estimates) or milk (RR 1.00, 95% CI 0.97–1.04, n=21 risk estimates) (**Figure 2** and Figure S6). Cheese intake was also associated with decreased risk of CHD and stroke (Figures S7 and S8). Considering the content of fat, consumption of low-fat dairy products was significantly related to lower total CVD (RR: 0.96, 95% CI: 0.92– 0.99, n=20 risk estimates) and stroke risk (RR: 0.90, 95% CI: 0.84–0.97, n=9 risk estimates) (**Figure 3** and Figure S9). No significant relationships were observed for high-fat dairy products (including high-fat milk, high-fat yogurt, high-fat cheese, and cream or butter) (**Figure 3** and Figure S10). For subtypes of stroke, milk consumption was related to a higher risk of hemorrhagic stroke (RR 1.08, 95% CI 1.01–1.17, n=5 risk estimates) and a decreased ischemic stroke risk was detected for total dairy (RR 0.92, 95% CI 0.86–0.99, n=7 risk estimates) and cheese consumption (RR 0.91, 95% CI 0.85–0.97, n=4 risk estimates) (Figures S11 to S14).

**Figure 2.**
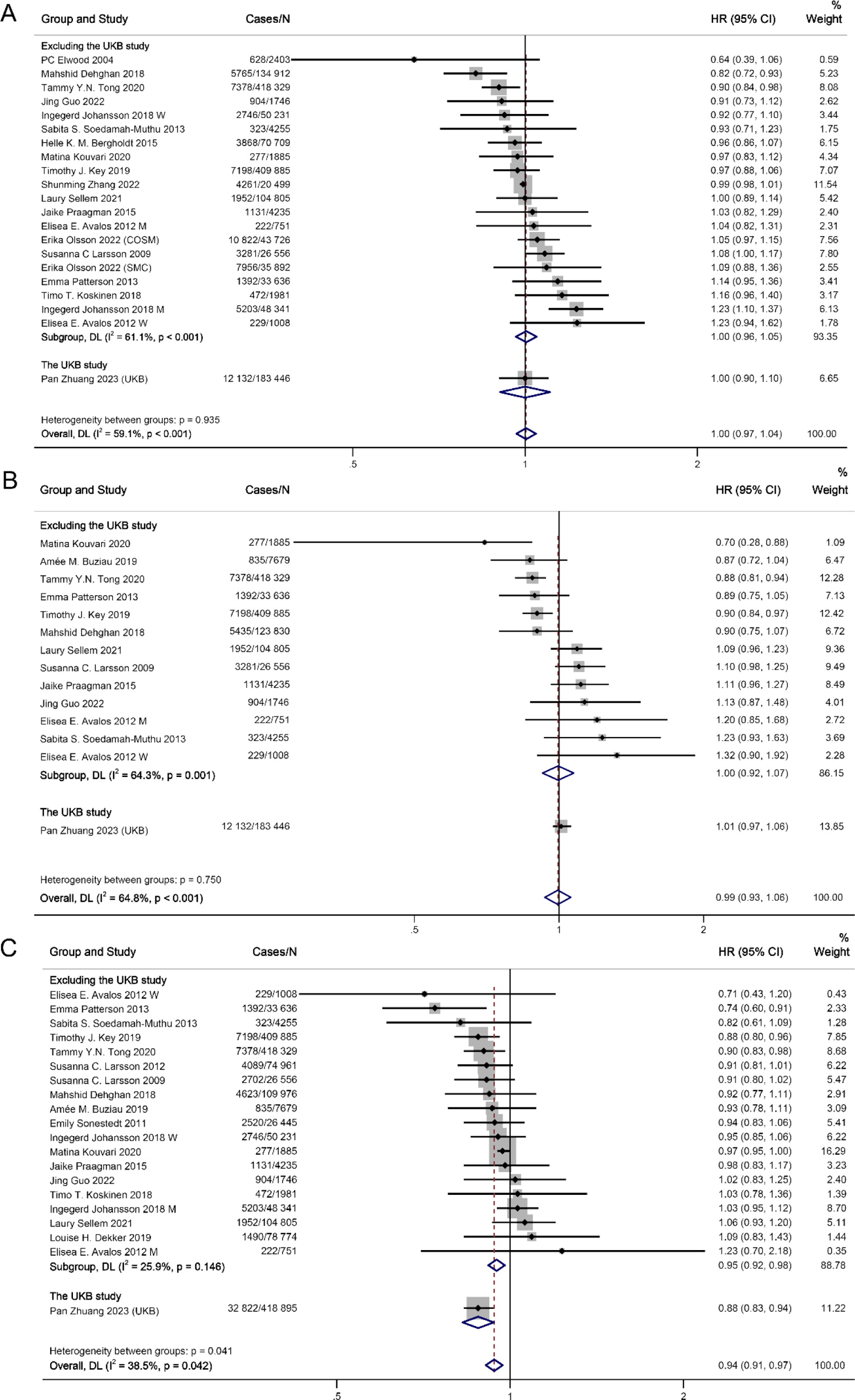
Associations of milk, yogurt, cheese consumption with cardiovascular disease risk for high compared with low category of intake using random effects meta-analysis. (A) Milk. (B) Yogurt. (C) Cheese. Squares represent study to Specific relative risk. Gray square areas are proportional to the individual study weight for the overall meta-analysis. Horizontal lines denote 95% CIs. I^2^ refers to the proportion of heterogeneity among studies. M, men; W, women; CKB, China Kadoorie Biobank; UKB, UK Biobank.

**Figure 3.**
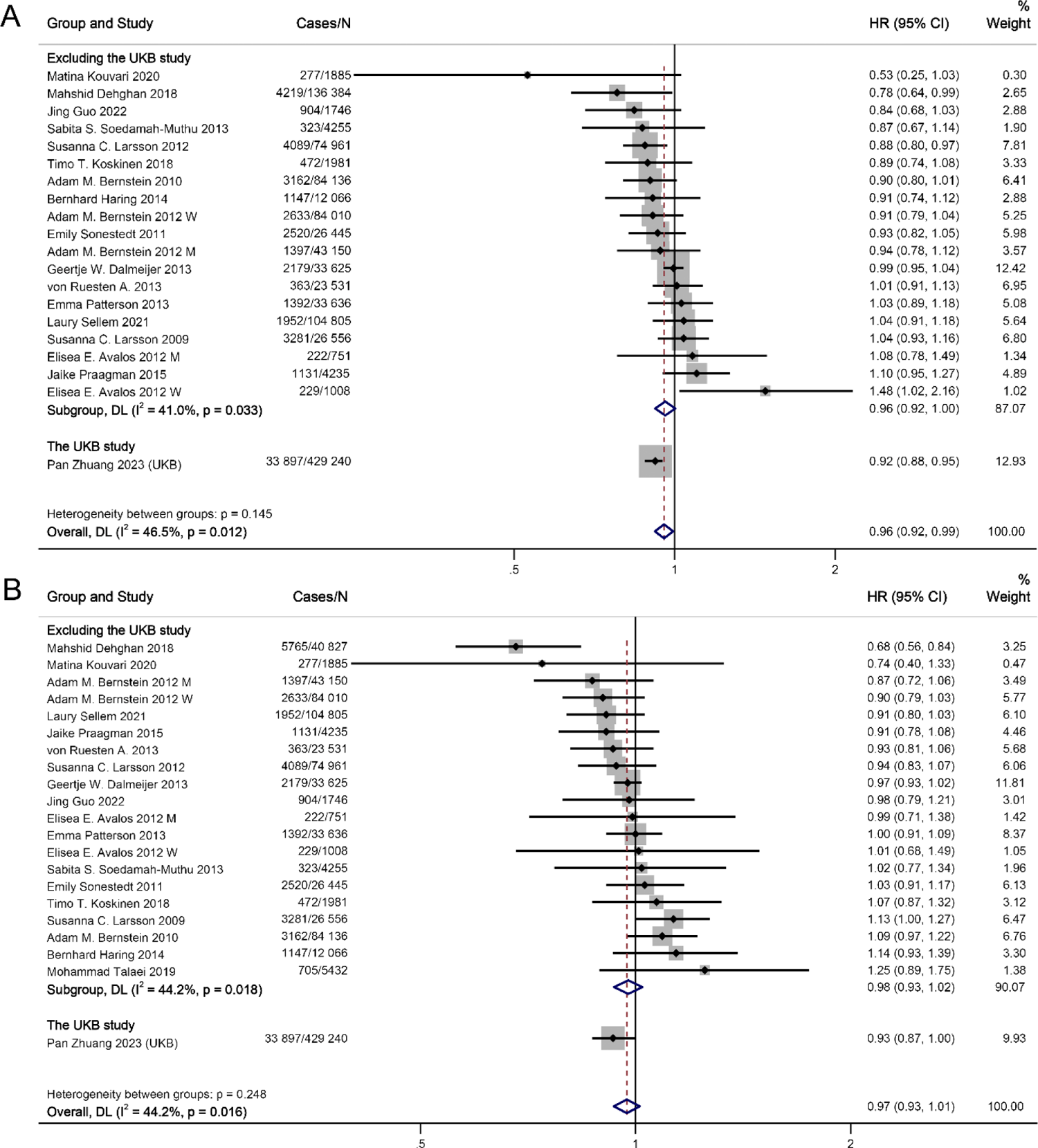
Associations of low-fat and high-fat dairy consumption with cardiovascular disease risk for high compared with low category of intake using random effects meta-analysis. (A) Low-fat. (B) High-fat. Squares represent study to Specific relative risk. Gray square areas are proportional to the individual study weight for the overall meta-analysis. Horizontal lines denote 95% CIs. I^2^ refers to the proportion of heterogeneity among studies. M, men; W, women; CKB, China Kadoorie Biobank; UKB, UK Biobank.

For total dairy consumption, we observed considerable heterogeneity across the studies (I^2^=66.1%) but did not find any publication bias (**Figure 1** and Figures S15 to S19). No significant heterogeneity was found in the predefined subgroup (sex, follow-up duration, region, Newcastle-Ottawa Scale score, etc.) meta-regressions (Table S26), indicating the source of heterogeneity mainly comes from subtypes of dairy. No single study disproportionately caused the heterogeneity (Figure S20).

Results of influence analysis for subtypes of dairy and subtypes of CVD are shown in Figures S21 to S30. If no significant heterogeneity was found across the studies for specific meta-analyses, we also conducted a fixed effects model to calculate summary HRs and 95% CIs which showed similar results (Table S27).

## DISCUSSION

In both UKB and CKB studies, dairy consumption was overall associated with lower risk of stroke. Further analysis of dairy subtypes in UKB revealed that cheese and skimmed/semi-skimmed milk consumption were inversely associated with CVD risk. The updated meta-analysis overall supported that dairy consumption, especially cheese and low-fat dairy consumption, was beneficial for CVD prevention among the general population.

### Comparison with Previous Studies and Possible Explanations

Our finding of the inverse association of dairy consumption with stroke risk was consistent with a recent meta-analysis showing a 1 to Serving/d increase in total dairy consumption was significantly related to a 4% decreased stroke risk.^20^ Although dairy products are major sources of saturated fatty acids (SFA) (about 65% of total fats), which has been shown to increase low-density lipoprotein (LDL) cholesterol levels, emerging evidence suggests that a low LDL cholesterol level (<70 mg/dL) was a risk factor for hemorrhagic stroke.^34, 35^ A meta-analysis summarizing data from 462,268 participants showed a dose-response relation of dietary SFA intake with lower stroke risk, especially intracranial hemorrhage risk.^36^ Congruously, we found that moderate intake of milk (0<milk intake≤0.5 serving/d) was associated with lower hemorrhagic stroke risk in UKB and total dairy consumption (mainly fresh milk/liquid whole milk in China)^37, 38^ was related to lower hemorrhagic stroke risk in CKB. Importantly, despite a high content of even-chain SFAs, dairy fats also consist of medium-chain (9.8%) and odd-chain (31.9%) SFAs,^39^ which may improve insulin sensitivity,^40^ Besides, dairy products also contain potentially beneficial natural trans fats, unsaturated fats, specific amino acids, branched-chain fats, vitamins K1 and K2, and calcium.^41^ Thus, given the complex food matrix of dairy products, their health impact cannot be fully accounted for by the presumed effect of SFAs. In our substitution analysis, replacing red meats with dairy products was related to lower stroke risk in CKB, reinforcing that the source of SFAs is more important. In addition, meta-analyses of randomized controlled trials demonstrated that fermented milk or dairy foods enriched with probiotics could reduce blood pressure,^42, 43^ which also partially explains the protective association for stroke, including a lower ischemic stroke risk for dairy in UKB and our meta-analysis.

With regard to CHD, we found great heterogeneity between UKB and CKB studies, which was also shown in our further updated meta-analysis (I^2^=68.6%). Further analyses revealed that the discrepancy between studies may be largely ascribed to different subtypes of dairy products. The protective relationship was mainly driven by cheese intake in the UKB study, which was further supported by our updated meta-analysis. Consistently, a meta-analysis of 15 prospective studies demonstrated that cheese consumption was related to reduced risk of CHD (RR [95% CI] for high vs. low consumption 0.86 [0.77–0.96]), stroke and total CVD.^44^ Another meta-analysis also showed a protective relationship of fermented dairy products with CVD risk and such protective association was detected for cheese but not yogurt.^19^

Although cheese, especially hard cheese, is rich in salt, saturated fat and calories, we still detected protective relationships for both hard cheese and fresh cheese in UKB. Potential mechanisms that underpin the relationship may be related to the high content of calcium, which may benefit cardiovascular health by limiting the absorption of SFAs and cholesterol^45^ and regulating the cell membrane potentials of the myocardium.^46^ Cheese also contains a high amount of conjugated linoleic acid that has been evidenced to inhibit the progression or induce the regression of atherosclerosis through modulating monocyte/macrophage function.^47^ In addition, the fermentation of dairy produced beneficial vitamin K_2_ that has been linked with a lower CHD risk.^48^ Microorganisms or probiotics from fermented dairy could modulate the gut microbiota composition, inhibit the reabsorption of bile acid, and produce beneficial short-chain fatty acids.^49^ A recent meta-analysis of 39 trials demonstrated that probiotic fermented milk products reduced serum total cholesterol and LDL cholesterol levels.^50^ However, our results of the updated meta-analysis and other meta-analyses found little benefit of yogurt consumption on CVD risk,^19, 51^ which could be due to the commonly added sugars or artificial sweeteners that might counteract the health benefit.^52^ Pertaining to milk consumption, mixed results have been reported from prospective studies.^19, 53^ A meta-analysis of cohort studies reported that milk intake was associated with a 4% (1%–5%) higher CHD mortality,^54^ which was congruent with our finding of a positive relation with CHD risk in CKB where liquid whole milk was the major dairy product.^37, 38^ In addition to the long even-chain SFAs elevating LDL cholesterol, a high D-galactose intake from non-fermented milk could be toxic.^55^ D-galactose has been widely used to establish an experimental model for premature aging by inducing oxidative stress and chronic inflammation,^55, 56^ which is also involved in the pathogenesis of CVD. Results from 2 large Swedish cohorts showed positive relations of milk intake with oxidative stress and inflammation markers while negative associations were observed for fermented milk products.^57^ Altogether, individual dairy products have divergent associations with CVD risk, which seemed to be the major reason for the discrepant results for CHD observed in CKB and UKB and also for the great heterogeneity between studies in our meta-analysis. Therefore, our study provides compelling evidence to highlight the importance of focusing on specific types of dairy products among which cheese may be the healthiest choice for the primary prevention of CVD.

Although prevailing dietary recommendations advocate consuming low-fat or non-fat dairy products over high-fat dairy/whole milk, previous evidence from meta-analyses showed no significant relations of low-fat dairy consumption with CVD or CHD risk.^19, 20^ Our meta-analysis showed inverse relationships of low-fat dairy consumption with CVD and stroke risk, supporting the protective role of low-fat dairy in CVD prevention. Nonetheless, we found no detrimental associations for high-fat dairy consumption. In a meta-analysis of 20 trials, both low-fat and high-fat dairy consumption increased body weight but had neutral effects on other cardiometabolic indicators, including waist circumference, fasting glucose, LDL cholesterol, high-density lipoprotein (HDL) cholesterol, blood pressure and C-reactive protein (CRP).^58^ Overall, current evidence suggests low-fat dairy may be beneficial for CVD whereas specific subtypes of high-fat dairy such as cheese could also be protective. More large studies are needed to compare low-fat with high-fat dairy on long-term CVD outcomes.

### Strengths and Limitations

This analysis has important strengths, including the large sample size, long follow-up duration, and the design of using data from two large cohorts in the UK and China, which enable us to directly compare the results from western vs. eastern countries. Finally, the updated meta-analysis provides a comprehensive overview of the evidence. Potential limitations also deserve attention. First, measurement errors by FFQs are inevitable in epidemiological studies. However, such errors tend to attenuate findings toward the null because of the prospective analysis. Although absolute dairy intake was not estimated in CKB and UKB, consumption frequency is rather useful in categorizing individuals on the basis of relative intakes. The consumption of subtypes of dairy products assessed from 24-hour recalls in UKB might not be representative of an individual’s long-term dietary habits. However, we calculated mean intakes from five separate occasions of 24-hour dietary recalls conducted from 2011 to 2012 (repeated measurement per person) to minimize this bias. Second, unmeasured or residual confounding cannot be fully ruled out despite our full adjustment for multiple risk factors. Specifically, higher dairy consumption seemed to be indicative of a higher socioeconomic status. Nonetheless, our results were consistent among both individuals with higher and lower income, indicating the documented associations of dairy were independent of socioeconomic status. Third, dairy consumption was measured once at baseline and changes over time were not considered. However, we still observed similar findings for a shorter follow-up duration (5 years), indicating that repeated measures are unlikely to affect our findings. Fourth, we could not further analyze dairy subtypes separately in CKB and butter was also not assessed in both CKB and UKB due to the lack of available data, which could have provided more implications.

## Conclusions and Implications

The results from our two large cohort studies and updated meta-analysis show that dairy consumption is associated with lower risk of stroke and total CVD overall while relationships for subtypes of dairy products differ. Cheese consumption but not milk or yogurt was inversely associated with CVD risk. Low-fat dairy consumption was inversely related to CVD and stroke risk. Our findings provide useful clinical evidence to advocate the consumption of dairy, especially cheese and low-fat dairy, for the primary prevention of CVD.

## Data Availability

The UK Biobank data are available from the UK Biobank on request (www.ukbiobank.ac.uk/). Details of how to access China Kadoorie Biobank data and details of the data release schedule are available from www.ckbiobank.org/site/Data+Access.

## Acknowledgements

We are grateful to UK Biobank and CKB participants. This research has been conducted using the UK Biobank resource (https://www.ukbiobank.ac.uk) under application number 47365 and CKB resource (https://www.ckbiobank.org/) under application number DAR-2020–00282. We thank Chinese Center for Disease Control and Prevention, Chinese Ministry of Health, National Health and Family Planning Commission of China, and 10 provincial/regional Health Administrative Departments. The most important acknowledgement is to the participants in the CKB study and the members of the survey teams in each of the 10 regional centers, as well as to the project development and management teams based at Beijing, Oxford and the 10 regional centers.

## Nonstandard Abbreviations and Acronyms

BMI: body mass index

CHD: coronary heart disease

CI: confidence interval

CKB: China Kadoorie Biobank

CRP: C-reactive protein

CVD: cardiovascular disease

FFQ: food frequency questionnaire

HDL: high-density lipoprotein

HR: hazard ratios

LDL: low-density lipoprotein

RR: relative risk

SFA: saturated fatty acids

TDI: Townsend deprivation index

UKB: UK Biobank.

